# Implications of the COVID-19 San Francisco Bay Area Shelter-in-Place Announcement: A Cross-Sectional Social Media Survey

**DOI:** 10.1101/2020.06.29.20143156

**Authors:** Holly Elser, Mathew V. Kiang, Esther M. John, Julia F. Simard, Melissa Bondy, Lorene M. Nelson, Wei-ting Chen, Eleni Linos

**Affiliations:** Stanford Medical School, Stanford University, CA, USA; Center for Population Health Sciences, Stanford University, CA, USA; Department of Epidemiology and Population Health, Stanford University, CA, USA; Office of Community Engagement, Stanford University, CA, USA; Department of Dermatology, Stanford University, CA, USA

## Abstract

**Background:** The U.S. has experienced an unprecedented number of shelter-in-place orders throughout the COVID-19 pandemic. There is limited empirical research that examines the impact of these orders. We aimed to rapidly ascertain whether social distancing; difficulty with daily activities (obtaining food, essential medications and childcare); and levels of concern regarding COVID-19 changed after the March 16, 2020 announcement of shelter-in-place orders for seven counties in the San Francisco Bay Area.

**Methods:** We conducted an online, cross-sectional social media survey from March 14 – April 1, 2020. We measured changes in social distancing behavior; experienced difficulties with daily activities (i.e., access to healthcare, childcare, obtaining essential food and medications); and level of concern regarding COVID-19 after the March 16 shelter-in-place announcement in the San Francisco Bay Area and elsewhere in the U.S.

**Results:** The percentage of respondents social distancing all of the time increased following the shelter-in-place announcement in the Bay Area (9.2%, 95% CI: 6.6, 11.9) and elsewhere in the U.S. (3.4%, 95% CI: 2.0, 5.0). Respondents also reported increased difficulty with obtaining food, hand sanitizer, and medications, particularly with obtaining food for both respondents from the Bay Area (13.3%, 95% CI: 10.4, 16.3) and elsewhere (8.2%, 95% CI: 6.6, 9.7). We found limited evidence that level of concern regarding the COVID-19 crisis changed following the shelter-in-place announcement.

**Conclusion:** These results capture early changes in attitudes, behaviors, and difficulties. Further research that specifically examines social, economic, and health impacts of COVID-19, especially among vulnerable populations, is urgently needed.

## Introduction

The coronavirus disease 2019 (COVID-19) pandemic began when clusters of “pneumonia of unknown etiology” were identified in December 2019. ^1-5^ By May 2020, there were over 3 million confirmed cases globally. One-third of these confirmed cases occurred in the United States (U.S.), with over 60,000 recorded deaths to date. ^6,7^

In the absence of vaccines or treatments, ^8^ the primary defense has been to reduce the risk of SARS-CoV-2 exposure through non-pharmaceutical interventions (NPIs) such as school closures, social distancing, isolation and quarantine, and use of personal masks. ^9-13^ NPIs were shown to be effective during the 2003 severe acute respiratory syndrome coronavirus (SARS-CoV) outbreak, ^14^ and quickly became the cornerstone of mitigation and intervention strategies for COVID-19 globally. ^15-17^ However, the extent and level of enforcement of these measures vary widely. ^9^

On March 19, 2020, California was the first U.S. state to enact a statewide shelter-in-place order, ^18^ following an announcement on March 16, 2020 of shelter-in-place orders for seven San Francisco Bay Area counties effective on 12:01 AM on March 17, 2020. ^19^ In the following weeks, 42 states and the District of Columbia passed shelter-in-place orders. ^20^ Potential subsequent SARS-CoV-2 wintertime outbreaks may necessitate repeated intermittent social distancing orders into 2022. ^17^ Given the unprecedented nature of these orders in the U.S., it is critical that we understand the impact of shelter-in-place orders on the public’s behaviors and perceptions.

For the present study, we employed convenience sampling to rapidly ascertain and summarize how levels of social distancing, difficulty related to daily activities such as obtaining food, essential medications and childcare, and levels of concern regarding the COVID-19 crisis changed after the March 16, 2020 shelter-in-place announcement among respondents living in the seven affected California counties compared with respondents living elsewhere in the U.S.

## Methods

### Study Sample

We conducted a cross-sectional, online survey with convenience sampling through three social media platforms (NextDoor, Twitter, and Facebook) starting on March 14, 2020 through April 1, 2020. Twitter and Facebook posts were shareable to facilitate snowball sampling. We included all respondents who completed at least 80% of the survey and excluded those missing both zip code and GeoIP location and those outside of the U.S.

### Data Collection

The 21-item survey collected information regarding shelter-in-place behaviors, experienced difficulty with daily activities, level of concern, demographic characteristics, and location. Demographic information included gender (female, male, other); race/ethnicity (white, Asian/ Pacific Islander, Hispanic/Latino, Black or other); year of birth was used to create age categories (25 years or less; 26 – 45; 46 – 65; older than 65 years); education (less than high school, high school or GED, some college, bachelor’s degree); and health insurance (yes, no, don’t know). Respondents reported the number of children (<18 years) and adults over age 65 years in their household. Participants were informed of the purpose, risks, and benefits of the study.

### Shelter-in-place announcement

We focused the analysis on the implications of shelter-in-place orders announced for six San Francisco Bay Area counties (San Francisco, Santa Clara, San Mateo, Marin, Contra Costa, and Alameda) and separately for Santa Cruz County made mid-day on March 16, 2020; hereafter, referred to collectively as “seven Bay Area counties”. We classified survey responses collected before March 16, 2020 as having occurred before the shelter-in-place announcement. We did so in order to more precisely identify responses that occurred before the announcement, as we anticipated that some respondents were aware of or suspected the shelter-in-place announcement several hours before it occurred. We differentiated survey respondents living in the seven affected Bay Area counties from those residing elsewhere in the U.S. using self-reported zip codes. For invalid or missing zip codes, we assigned participants’ locations based on latitude and longitude (i.e., GeoIP location, an estimation of the respondent’s location based on their IP address).

### Level of concern, social distancing behaviors, and difficulties

We considered three outcomes: social distancing behaviors (all of the time, most of the time, some of the time, none of the time); experienced difficulties with daily activities (access to healthcare, childcare, transportation, job loss, or difficulty obtaining essential items including food, medications, and hand sanitizer); and level of concern regarding the COVID-19 crisis (extremely concerned, very concerned, moderately concerned, somewhat concerned, not at all concerned).

### Statistical Analysis

We first summarized demographic characteristics for survey respondents living in the seven Bay Area counties affected by the shelter-in-place announcement on March 16, 2020 compared to respondents living elsewhere within the U.S.

#### Changes Before and After the Shelter-in-Place Announcement

We used linear probability models to assess changes in levels of social distancing, the proportion of respondents experiencing difficulty with daily activities, and level of concern regarding the COVID-19 crisis after versus before the shelter-in-place announcement separately for respondents in the seven Bay Area counties and for respondents elsewhere in the U.S. Beta coefficients were transformed to reflect percent changes in each response level after the announcement was made.

#### Difference-in-Differences Estimates

We used a difference-in-differences (DID) approach with linear probability models to estimate the impact of the shelter-in-place announcement. ^21,22^ The DID estimator compared the change in responses after versus before March 16, 2020 among respondents in the Bay Area versus elsewhere in the U.S. The DID approach assumes that any changes that occurred outside of the Bay Area reflect background or secular trends. Under the assumption that these trends would have been parallel among respondents in the Bay Area and elsewhere had the shelter-in-place announcement not occurred, the resulting DID estimates correspond to the change in each outcome attributable to the announcement itself in the Bay Area. We calculated DID estimates in the study population overall, and within subgroups defined by gender, age, and household composition (at least one child at home, at least one adult > 65 years).

#### Sensitivity Analyses

We considered three alternative approaches for the DID analysis. First, we compared responses from the entire state of California to those of respondents elsewhere in the U.S. Because the announcement was highly publicized on mainstream news media channels and social media platforms, survey respondents living in California outside of the seven Bay Area counties may have modified their behaviors. We therefore repeated the main analysis comparing respondents in California to respondents in other U.S. states. Second, a similar announcement also occurred in Washington state on March 16, 2020. Therefore, we repeated the main analysis with respondents from Washington state combined with Bay Area respondents. Finally, we repeated the main analysis excluding responses after March 19, 2020 – when California announced a state-wide shelter-in-place order.

We conducted all statistical analyses using R version 3.2.3 (R Foundation for Statistical Computing, Vienna, Austria). This study was approved by the Institutional Review Board at Stanford University.

## Results

In total, 22,913 respondents started the survey. We excluded 4,031 respondents who completed less than 80% of the survey, 1,136 respondents with no geolocation data, and 203 international respondents. The final analytic sample included 17,543 respondents of whom 4,161 (24%) were from the seven Bay Area counties. Among respondents from the Bay Area, 2,951 (70.9%) completed the survey prior to March 16, 2020. Among respondents living elsewhere in the U.S., 8,410 (62.8%) completed the survey prior to March 16, 2020 (**Table 1**). Overall, the majority of respondents were younger than 66 years (N = 90%), and the majority (84%) had earned at least a bachelor’s degree. The majority of respondents were female (72%), and most (96%) had some form of health insurance. Approximately 41% of respondents indicated living with at least one child under the age of 18 years and 19% indicated living with at least one adult over the age of 65 years.

**Table 1.**
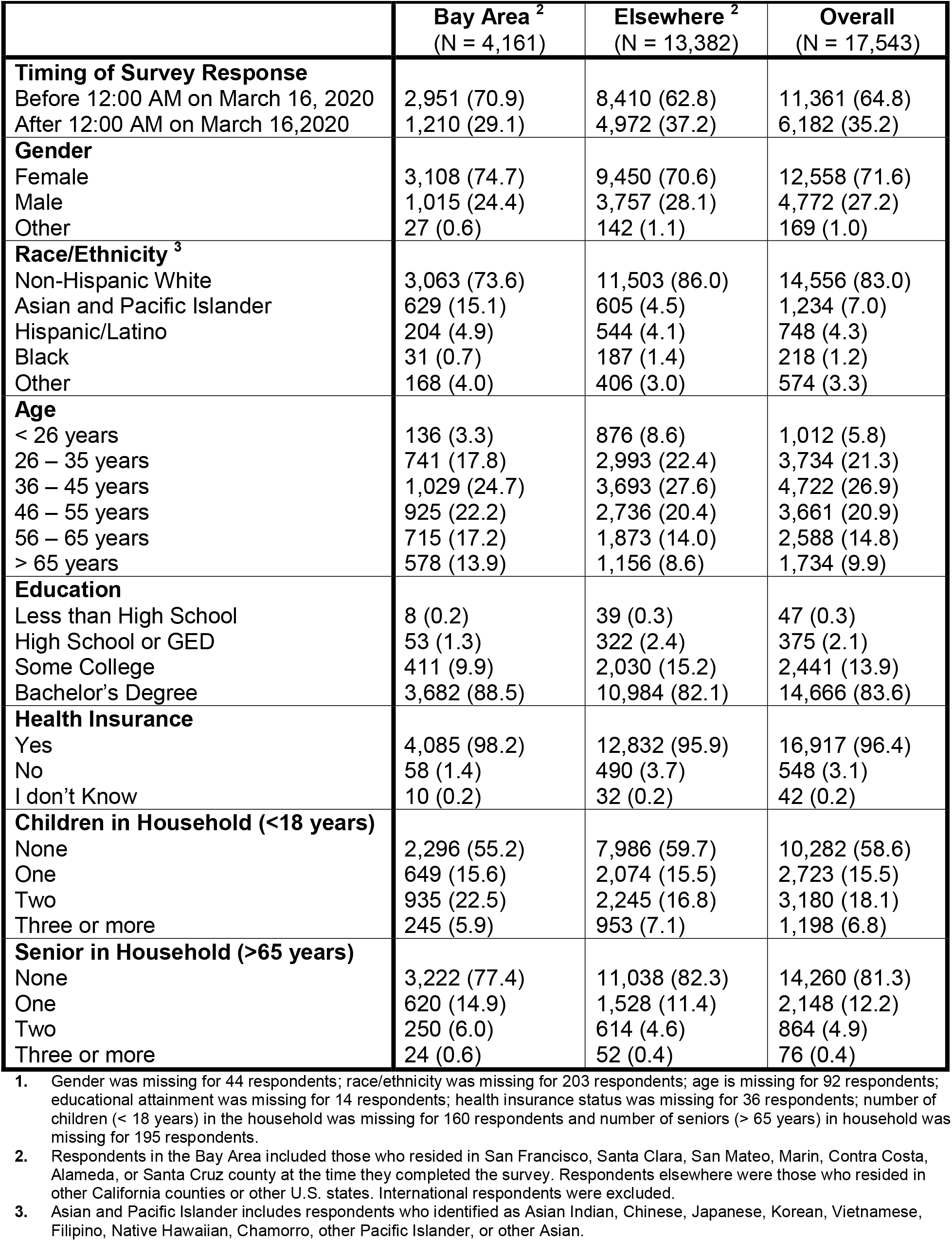
Demographic characteristics for California and in the study population overall – N (%) ^**1**^

|  | **Bay Area <sup>2</sup>**<br>(N = 4,161) | **Elsewhere <sup>2</sup>**<br>(N = 13,382) | **Overall**<br>(N = 17,543) |
| --- | --- | --- | --- |
| **Timing of Survey Response** |  |  |  |
| Before 12:00 AM on March 16, 2020 | 2,951 (70.9) | 8,410 (62.8) | 11,361 (64.8) |
| After 12:00 AM on March 16,2020 | 1,210 (29.1) | 4,972 (37.2) | 6,182 (35.2) |
| **Gender** |  |  |  |
| Female | 3,108 (74.7) | 9,450 (70.6) | 12,558 (71.6) |
| Male | 1,015 (24.4) | 3,757 (28.1) | 4,772 (27.2) |
| Other | 27 (0.6) | 142 (1.1) | 169 (1.0) |
| **Race/Ethnicity <sup>3</sup>** |  |  |  |
| Non-Hispanic White | 3,063 (73.6) | 11,503 (86.0) | 14,556 (83.0) |
| Asian and Pacific Islander | 629 (15.1) | 605 (4.5) | 1,234 (7.0) |
| Hispanic/Latino | 204 (4.9) | 544 (4.1) | 748 (4.3) |
| Black | 31 (0.7) | 187 (1.4) | 218 (1.2) |
| Other | 168 (4.0) | 406 (3.0) | 574 (3.3) |
| **Age** |  |  |  |
| < 26 years | 136 (3.3) | 876 (8.6) | 1,012 (5.8) |
| 26 – 35 years | 741 (17.8) | 2,993 (22.4) | 3,734 (21.3) |
| 36 – 45 years | 1,029 (24.7) | 3,693 (27.6) | 4,722 (26.9) |
| 46 – 55 years | 925 (22.2) | 2,736 (20.4) | 3,661 (20.9) |
| 56 – 65 years | 715 (17.2) | 1,873 (14.0) | 2,588 (14.8) |
| > 65 years | 578 (13.9) | 1,156 (8.6) | 1,734 (9.9) |
| **Education** |  |  |  |
| Less than High School | 8 (0.2) | 39 (0.3) | 47 (0.3) |
| High School or GED | 53 (1.3) | 322 (2.4) | 375 (2.1) |
| Some College | 411 (9.9) | 2,030 (15.2) | 2,441 (13.9) |
| Bachelor's Degree | 3,682 (88.5) | 10,984 (82.1) | 14,666 (83.6) |
| **Health Insurance** |  |  |  |
| Yes | 4,085 (98.2) | 12,832 (95.9) | 16,917 (96.4) |
| No | 58 (1.4) | 490 (3.7) | 548 (3.1) |
| I don't Know | 10 (0.2) | 32 (0.2) | 42 (0.2) |
| **Children in Household (<18 years)** |  |  |  |
| None | 2,296 (55.2) | 7,986 (59.7) | 10,282 (58.6) |
| One | 649 (15.6) | 2,074 (15.5) | 2,723 (15.5) |
| Two | 935 (22.5) | 2,245 (16.8) | 3,180 (18.1) |
| Three or more | 245 (5.9) | 953 (7.1) | 1,198 (6.8) |
| **Senior in Household (>65 years)** |  |  |  |
| None | 3,222 (77.4) | 11,038 (82.3) | 14,260 (81.3) |
| One | 620 (14.9) | 1,528 (11.4) | 2,148 (12.2) |
| Two | 250 (6.0) | 614 (4.6) | 864 (4.9) |
| Three or more | 24 (0.6) | 52 (0.4) | 76 (0.4) |

1. Gender was missing for 44 respondents; race/ethnicity was missing for 203 respondents; age is missing for 92 respondents; educational attainment was missing for 14 respondents; health insurance status was missing for 36 respondents; number of children (< 18 years) in the household was missing for 160 respondents and number of seniors (> 65 years) in household was missing for 195 respondents. 2. Respondents in the Bay Area included those who resided in San Francisco, Santa Clara, San Mateo, Marin, Contra Costa, Alameda, or Santa Cruz county at the time they completed the survey. Respondents elsewhere were those who resided in other California counties or other U.S. states. International respondents were excluded. 3. Asian and Pacific Islander includes respondents who identified as Asian Indian, Chinese, Japanese, Korean, Vietnamese, Filipino, Native Hawaiian, Chamorro, other Pacific Islander, or other Asian.

Respondents from the Bay Area were less likely to identify as non-Hispanic white as compared with other respondents (73.6% versus 86.0%) and less likely to identify as Black (0.7% versus 1.4%). Respondents from the Bay Area were more likely to be Asian or Pacific Islanders (15.1% versus 4.5%) or Hispanic/Latino (4.9% versus 4.1%). Respondents from the Bay Area were also less likely to be under age 36 years (21.1% versus 31.0%) and slightly more likely to be over age 65 years (13.9% versus 8.6%). The distribution of participants by gender, educational attainment, and household composition was similar among respondents from the Bay Area and respondents living elsewhere. We noted only minor differences between respondents who completed the survey before or after March 16, 2020 in the Bay Area or elsewhere, except for the percentage of respondents who were female and living outside of the Bay Area which was substantially lower before March 16, 2020 versus afterwards (52.5% versus 79.1%). (**Table A1 in the Supplementary Materials**)

### Changes Before and After the Shelter-in-Place Announcement

In **Table 2**, we present the change in level of social distancing, difficulties experienced, and level of concern following the March 16, 2020 shelter-in-place announcement for respondents from the Bay Area and respondents living elsewhere. In general, we observed similar trends in the two groups. We found an increase in the proportion of respondents practicing social distancing all of the time after the shelter-in-place announcement in the Bay Area (9.2%, 95% CI: 6.6, 11.9) and elsewhere (3.4%, 95% CI: 2.0, 5.0). We also observed increases in the proportion sheltering in place most of the time among survey respondents from the Bay Area (5.7%, 95% CI: 2.3, 9.0) and elsewhere (8.5%, 95% CI: 6.8, 10.3). The proportion of respondents sheltering in place some of the time and none of the time decreased both among respondents from the Bay Area and elsewhere.

**Table 2.** Changes in social distancing, difficulties, and concern after the shelter-in-place versus before in the Bay Area versus elsewhere in the U.S.

|  | Bay Area |  |  | Elsewhere |  |  |
| --- | --- | --- | --- | --- | --- | --- |
|  | Before – %<br>(N = 2,951) | After – %<br>(N = 1,210) | Percent Change<br>(95% CI) | Before – %<br>(N = 8,410) | After – %<br>(N = 4,972) | Percent Change<br>(95% CI) |
| **Social Distancing**<sup>1</sup> |  |  |  |  |  |  |
| All of the time | 17.3 | 26.5 | 9.21 (6.55, 11.9) | 19.1 | 22.6 | 3.40 (1.99, 4.81) |
| Most of the time | 54.4 | 60.0 | 5.65 (2.33, 8.96) | 48.1 | 56.7 | 8.54 (6.79, 10.3) |
| Some of the time | 26.7 | 12.3 | 1.41 (- 17.2, - 11.6) | 29.4 | 18.6 | - 10.9 (- 12.3, - 9.26) |
| None of the time | 1.6 | 1.2 | - 4.70 (- 5.51, - 3.88) | 3.3 | 2.2 | - 1.11 (- 1.70, - 0.53) |
| **Difficulties**<sup>2</sup> |  |  |  |  |  |  |
| Access to Healthcare | 4.2 | 7.4 | 3.19 (1.72, 4.66) | 4.2 | 7.0 | 2.83 (2.05, 3.60) |
| Childcare | 15.1 | 15.3 | 0.18 (- 2.22, 2.58) | 9.4 | 13.1 | 3.61 (2.53, 4.70) |
| Food | 23.8 | 37.1 | 13.3 (10.4, 16.3) | 23.5 | 31.6 | 8.17 (6.63, 9.71) |
| Job Loss | 0.6 | 1.8 | 1.17 (0.51, 1.83) | 1.4 | 2.9 | 1.56 (1.07, 2.04) |
| Medications | 7.6 | 8.9 | 0.93 (- 0.48, 3.15) | 7.1 | 8.6 | 1.52 (0.59, 2.45) |
| Sanitizer | 63.1 | 68.8 | 5.71 (2.52, 8.90) | 59.0 | 62.5 | 3.50 (1.78, 5.21) |
| Transportation | 2.8 | 4.7 | 1.93 (0.73, 3.13) | 3.2 | 3.7 | 0.55 (- 0.08, 1.18) |
| Wages | 9.4 | 14.1 | 4.70 (2.63, 6.76) | 11.3 | 17.7 | 6.41 (5.22, 7.61) |
| **Level of Concern**<sup>3</sup> |  |  |  |  |  |  |
| Extremely concerned | 29.0 | 28.2 | - 0.83 (- 3.85, 2.20) | 32.9 | 28.9 | - 4.05 (- 5.68, - 2.43) |
| Very concerned | 36.9 | 36.1 | - 0.75 (- 3.98, 2.47) | 35.0 | 36.8 | 1.79 (0.11, 3.47) |
| Moderately concerned | 25.6 | 26.3 | 0.73 (- 2.20, 3.66) | 23.1 | 24.8 | 1.75 (0.26, 3.24) |
| A little concerned | 7.5 | 8.2 | 0.73 (- 1.05, 2.51) | 7.4 | 8.1 | 0.74 (- 0.19, 1.67) |
| Not at all concerned | 1.1 | 1.2 | 0.12 (- 0.59, 0.84) | 1.6 | 1.4 | - 0.24 (- 0.67, 0.19) |

1. Respondents were asked to select their level of social distancing. We created a mutually exclusive set of indicator variables.
2. Respondents were asked to select all of the difficulties they had experienced because of the COVID-19 crisis; categories are not mutually exclusive.
3. Respondents were asked to select their level of concern regarding the COVID-19 crisis. We created a mutually exclusive set of indicator variables.
4. We used generalized linear probability models to estimate the change in level of concern, social distancing levels, and experienced difficulties before and after the announcement. We calculated percentages by multiplying coefficients by 100.

Respondents also reported more difficulty associated with activities such as obtaining food, hand sanitizer, and medications after the March 16, 2020 shelter-in-place announcement versus before. The increase in difficulty was largest for obtaining food for both respondents from the Bay Area (13.3%, 95% CI: 10.4, 16.3) and elsewhere (8.2%, 95% CI: 6.6, 9.7). Similarly, both groups reported greater difficulty obtaining hand sanitizer. Greater difficulty with wages was reported more frequently by respondents from the Bay Area following the shelter-in-place announcement (4.7%, 95% CI: 2.6, 6.8) and even more so by respondents living elsewhere (6.4%, 95% CI: 5.2, 7.6). Respondents in both groups were also more likely to report difficulty related to job loss following the announcement (Bay Area: 1.2%, 95% CI: 0.5, 1.8; Elsewhere: 1.6%, 95% CI 1.1, 2.0).

We observed only small changes in level of concern regarding the COVID-19 crisis after the March 16, 2020 shelter-in-place announcement among respondents in the Bay Area. Among respondents living elsewhere, we observed a decrease in the proportion of respondents reporting they were “extremely concerned” after the announcement (−4.1%, 95% CI: - 5.7, - 2.4).

### Difference-in-Differences Estimates

In **Table 3**, we present DID estimates for the change in the proportion of respondents who were social distancing all of the time after the shelter-in-place announcement in the Bay Area versus elsewhere.

**Table 3.** Percentage of respondents who were social distancing all of the time in Bay Area versus elsewhere in the U.S. before and after the March 16^th^, 2020 Bay Area Shelter-in-Place Announcement and difference-in-differences estimates for the study population overall and within strata of gender, age category, and household composition. ^1^

|  | Bay Area |  | Elsewhere |  | DID Estimate<br>(95% CI) |
| --- | --- | --- | --- | --- | --- |
|  | Before – %<br>(N = 2,951) | After – %<br>(N = 1,210) | Before – %<br>(N = 8,410) | After – %<br>(N = 4,972) |  |
| **Overall** <sup>2</sup> | 511 (17.3) | 321 (26.5) | 1,610 (19.1) | 1,121 (22.5) | 5.81 (2.78, 8.84) |
| **Sex** <sup>3</sup> |  |  |  |  |  |
| Women | 392 (18.0) | 244 (26.3) | 1,083 (19.6) | 915 (23.3) | 4.62 (1.08, 8.16) |
| Men | 113 (15.1) | 70 (26.1) | 501 (18.1) | 194 (19.7) | 9.32 (3.22, 15.4) |
| **Age Category** <sup>4</sup> |  |  |  |  |  |
| 25 Years or Less | 14 (16.7) | 10 (19.2) | 69 (11.5) | 42 (15.2) | - 1.15 (- 13.9, 11.6) |
| 26 – 45 Years | 217 (18.0) | 149 (26.3) | 782 (19.0) | 577 (22.5) | 4.72 (0.24, 9.20) |
| 46 – 65 Years | 181 (14.9) | 96 (22.4) | 608 (20.1) | 330 (20.9) | 6.72 (1.75, 11.7) |
| Older than 65 Years | 91 (21.5) | 65 (41.9) | 149 (23.7) | 168 (31.9) | 12.1 (2.53, 21.7) |
| **Household Composition** <sup>5</sup> |  |  |  |  |  |
| Households with child < 18 | 240 (17.5) | 132 (29.1) | 679 (20.5) | 448 (23.2) | 8.99 (4.09, 13.9) |
| Households with adult > 65 | 115 (18.3) | 80 (30.2) | 306 (21.9) | 222 (27.9) | 5.91 (- 1.18, 13.0) |

1. We used a difference-in-difference in estimator that compared the change in response following the March 16, 2020 shelter-in-place announcement in California versus elsewhere. We calculated the percent change in California vs. elsewhere by multiplying linear probability estimates by 100. 2. Percentages and DID estimate for the study population overall (N = 17,543) 3. Percentages and DID estimates for subgroup of women (N = 12,558) and men (N = 4,772) 4. Percentages and DID estimates among respondents less than 25 years old (N = 744), between the ages 25 and 34 (N = 3,493), between the ages of 35 and 44 (N = 4,807), between the ages of 45 and 54 (N = 3,790), between the ages of 55 and 64 (N = 2,679) and 65 years or older (N = 1,938) 5. Percentages and DID estimates for household with at least one child (N = 7,261) and at least one elderly household member (N = 3,283)

Overall, the proportion of respondents social distancing all of the time increased after the announcement in the Bay Area versus elsewhere (5.8%, 95% CI: 2.8, 8.8). Relative increases were greatest among men (9.3%, 95% CI: 3.2, 15.4), adults between the ages of 46 and 65 years (6.7%, 95% CI 1.8, 11.7), and respondents from households with children.

We calculated DID estimates for experienced difficulties in the Bay Area versus elsewhere following the shelter-in-place announcement. We noted the strongest differences for difficulty obtaining food (5.2%, 95% CI: 1.8, 8.5), followed by difficulty with transportation (2.2, 95% CI: - 1.5, 5.9) (**Figure 1, Table A2**). We observed limited evidence of increased difficulty with healthcare, obtaining hand sanitizer, or obtaining medications.

**Figure 1.**
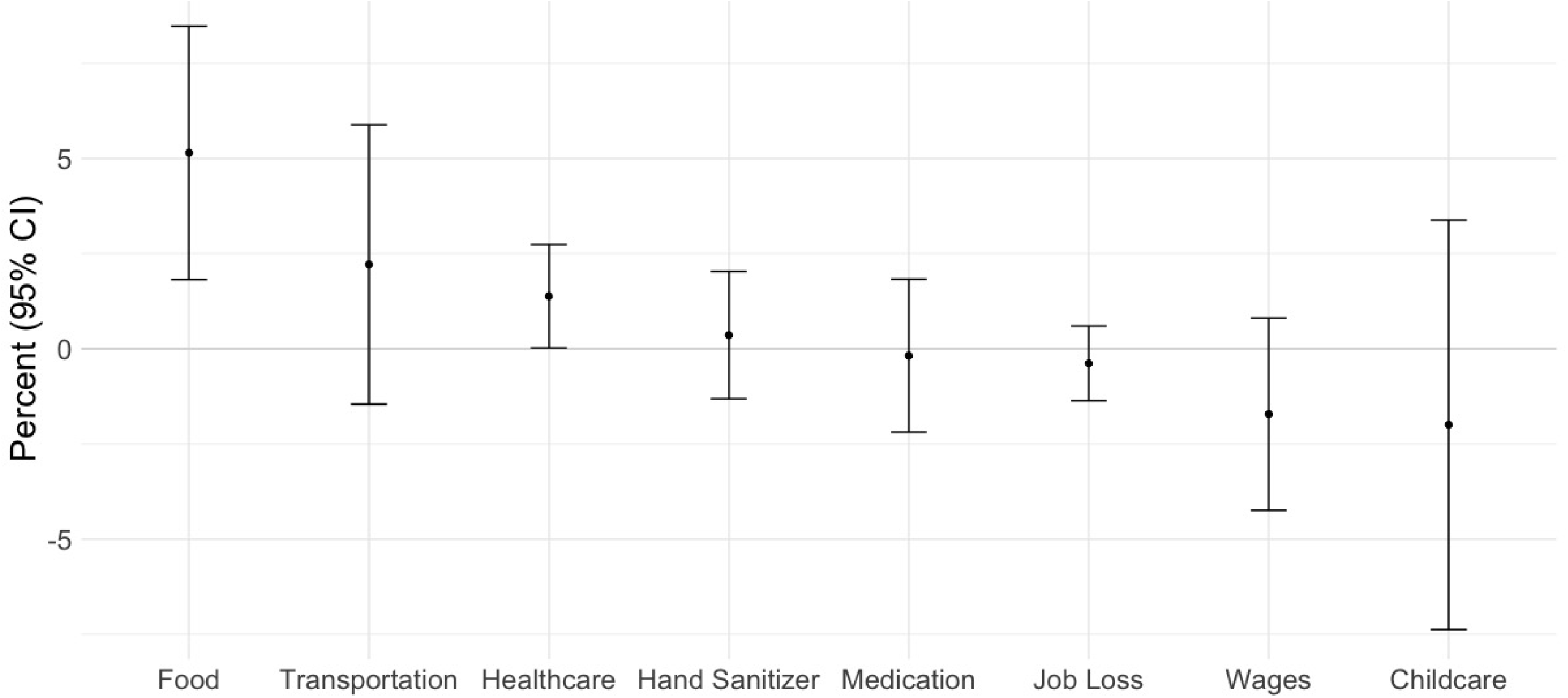
Difference-in-difference estimates for experienced difficulties in California versus elsewhere following the March 16, 2020 announcement of the Bay Area shelter in place order. We used linear probability models to estimate the change in California versus elsewhere for each of the above experienced difficulties for the full sample (N = 17,543) and among the subset of a respondents living in a household with a child < 18 for difficulty with childcare (N = 7,062). We transformed model coefficients into percentages by multiplying estimated proportions by 100%

In **Table 4** we present DID estimates for the change in the proportion of respondents who were extremely concerned following the shelter-in-place announcement in the Bay Area versus elsewhere. Overall, the proportion of respondents who reported extreme worry did not increase after the announcement for most groups, with the exception of those aged 46-65 years (8.03, 95% CI 2.03, 14.0) and respondents living with at least one child (6.20, 95% CI 0.62, 11.8). The proportion reporting extreme worry decreased in some groups including men, those under age 25, and those living outside the Bay Area.

**Table 4.** Percentage of respondents who were extremely worried about the COVID-19 crisis in the Bay Area and elsewhere in the U.S. before and after the March 16^th^, 2020 Bay Area Shelter-in-Place Announcement and difference-in-differences estimates for the study population overall and within strata of gender, age category, and household composition. ^1^

|  | Bay Area |  | Elsewhere |  | DID Estimate<br>(95% CI) |
| --- | --- | --- | --- | --- | --- |
|  | Before – %<br>(N = 2,951) | After – %<br>(N = 1,210) | Before – %<br>(N = 8,410) | After – %<br>(N = 4,972) |  |
| **Overall** <sup>2</sup> | 856 (29.0) | 341 (28.2) | 2,768 (32.9) | 1,435 (28.9) | 3.23 (- 0.26, 6.71) |
| **Sex** <sup>3</sup> |  |  |  |  |  |
| Women | 643 (29.5) | 280 (30.1) | 1,880 (34.1) | 1,204 (30.6) | 4.07 (0.01, 8.12) |
| Men | 205 (27.4) | 55 (20.5) | 851 (30.7) | 218 (22.2) | 1.58 (- 5.45, 8.61) |
| **Age Category** <sup>4</sup> |  |  |  |  |  |
| 25 Years or Less | 13 (15.5) | 6 (11.5) | 124 (20.7) | 39 (14.1) | 2.60 (- 1.17, 16.9) |
| 26 – 45 Years | 267 (22.2) | 146 (25.7) | 1,281 (31.1) | 673 (26.2) | 0.09 (- 4.97, 5.15) |
| 46 – 65 Years | 355 (29.3) | 148 (34.6) | 1,143 (37.8) | 554 (35.0) | 8.03 (2.03, 14.0) |
| Older than 65 Years | 113 (26.7) | 40 (25.8) | 210 (33.3) | 161 (30.6) | 1.82 (- 8.16, 11.8) |
| **Household Composition** <sup>5</sup> |  |  |  |  |  |
| Households with child < 18 | 421 (30.6) | 146 (32.2) | 1,105 (33.4) | 556 (28.8) | 6.20 (0.62, 11.8) |
| Households with adult > 65 | 179 (28.5) | 74 (27.9) | 511 (33.6) | 268 (33.7) | 2.35 (- 5.55, 10.3) |

1. We used a difference-in-difference in estimator that compared the change in response following the March 16, 2020 shelter-in-place announcement in California versus elsewhere. We calculated the percent change in California vs. elsewhere by multiplying linear probability estimates by 100. 2. Percentages and DID estimates for the study population overall (N = 17,543) 3. Percentages and DID estimates for subgroup of women (N = 12,558) and men (N = 4,772) 4. Percentages and DID estimates among respondents less than 25 years old (N = 744), between the ages 25 and 34 (N = 3,493), between the ages of 35 and 44 (N = 4,807), between the ages of 45 and 54 (N = 3,790), between the ages of 55 and 64 (N = 2,679) and 65 years or older (N = 1,938) 5. Percentages and DID estimates for household with at least one child (N = 7,261) and at least one elderly household member (N = 3,283)

### Sensitivity Analyses

The overall pattern of results in all three sensitivity analyses was consistent with our main analysis. Effect estimates were slightly attenuated when we compared California respondents to respondents elsewhere in the U.S., or when we combined Washington state respondents with Bay Area respondents (Supplemental **tables A3, A4, A5**), and slightly accentuated when we excluded survey responses after March 19, 2020.

## Discussion

We examined changes in attitudes and behaviors as a result of the COVID-19 crisis in a cross-sectional convenience sample of 17,543 respondents recruited through three social media platforms. Differences in key demographic characteristics (level of insurance, educational attainment, race/ethnicity) preclude generalization of our findings to the Bay Area or to the U.S. more broadly. Nevertheless, the study data capture how social distancing behaviors, difficulties with daily activities, and levels of concern regarding how COVID-19 may have changed in the days that immediately preceded and immediately followed the announcement of the nation’s first shelter-in-place order for seven Bay Area counties. This announcement occurred at a point where the seriousness of the COVID-19 pandemic for the U.S. was increasingly recognized, but the eventual impact on cities such as New York, New Orleans, and Detroit was yet to be realized. ^23-26^ As such, the results of this study offer some insight into our collective disposition towards the pandemic at a unique point in history as the very first shelter in place policy decisions were made.

Overall, we found that participants’ behaviors and attitudes regarding the COVID-19 pandemic evolved even within our brief survey period. After the shelter-in-place orders were announced for the Bay Area, social distancing increased. Increases in level of social distancing were more pronounced among respondents in the Bay Area versus those living elsewhere in the U.S., adults older than 46 years, and those living with children or an adult over age 65 years. This pattern may be explained by early suspicions that older adults were most vulnerable to COVID-19. ^27^

Consistent with media reports of food shortages, ^28^ respondents were most likely to report difficulty obtaining food, with increases in difficulty obtaining food more pronounced in the Bay Area following the shelter-in-place announcement. Increases in difficulty with access to healthcare, hand sanitizer, and transportation were similar among respondents in the Bay Area versus those living elsewhere. We detected the early impacts on job loss and wages, which were followed by a national surge in unemployment after the study period. ^29,30^ We anticipate that our findings may further underestimate the impacts of shelter-in-place on job loss and wages given the high levels of educational attainment in our study population, as may respondents may have been able to transition more easily to remote work. ^31^

Finally, we found that approximately one-third of respondents were “extremely concerned” about the COVID-19 crisis, although we found little evidence to support the idea that levels of concern increased – among respondents in the Bay Area or elsewhere – following the announcement of shelter-in-place orders. This raises the interesting question as to whether announcements regarding COVID-19 lead to increased or decreased levels of concern and anxiety that should be considered further in more representative study populations and as the pandemic continues to evolve.

### Limitations

Despite the large number of survey respondents, older adults, Black respondents, and men were underrepresented in this convenience sample. Similarly, household structure of respondents suggests that a large number of respondents did not have children or elderly family members that may have required extra care. Recruitment was convenience sampling via three social media websites. Snowball sampling (through re-posts on Facebook and Twitter) may have further propagated participation among a more homogenous group of respondents. Our results therefore likely underrepresent the true extent of challenges associated with the pandemic across the U.S. and precludes meaningful examination of the early impacts of shelter-in-place orders on economically marginalized and vulnerable population subgroups. ^32,33^

The cross-sectional nature of this study represents an additional limitation. Because we did not observe changes in social distancing, experienced difficulties, and levels of concern in individuals over time, it is possible that our findings are explained at least in part by compositional effects (i.e., systematic differences in respondents who completed the survey before and after March 16^th^). Reassuringly, we found limited evidence of systematic differences in measured characteristics before and after the March 16^th^ cutoff with the exception of the gender breakdown among respondents who resided outside of the Bay Area.

Finally, the announcement of shelter-in-place orders for the seven Bay Area counties was covered extensively in the national media, which makes spillover effects of the announcement to survey respondents living outside of the Bay Area – particularly elsewhere in California –likely. The assumptions of DID are therefore unlikely to be met, and our estimates are more appropriately interpreted as summary measures of the change in the Bay Area relative to the change elsewhere in the U.S. rather than causal estimates of the impact of the announcement. However, in sensitivity analyses to examine spillover in Washington State and California, we found similar pattern of findings across subgroups of interest.

## Conclusions

We found evidence of increased social distancing and difficulty with daily activities such as food and transportation in the wake of the announcement of the nation’s first shelter-in-place orders, particularly among respondents in the Bay Area. Levels of concern remained fairly consistent throughout the study period among respondents in the Bay Area and elsewhere. Given that our study population was highly educated, concentrated in one of the more affluent areas in the U.S., and queried relatively early in the COVID-19 pandemic, we anticipate that our findings underestimate substantially the impact of county- and statewide shelter-in-place orders. As such, our study represents a first step towards understanding the social attitudes and consequences of this crisis. Further research that specifically examines social, economic, and health impacts of COVID-19 especially among vulnerable populations is needed.

## Data Availability

Data will be provided upon request.

